# Mechanical Instability as a Signature of Viscoelastic Decoupling at the Tumor–Brain Interface

**DOI:** 10.1101/2025.11.15.25340289

**Authors:** Jan Saip Aunan-Diop, Ancuta Ioana Friismose, Emi Hojo, Yin Ziying, Bo Halle, Frederik Harbo, Bo Mussmann, Frantz Rom Poulsen

## Abstract

Brain tumors alter the viscoelastic equilibrium of surrounding tissue, but how these changes shape the mechanics of tumor–brain coupling remains unclear. This study introduces mechanical instability mapping, a voxelwise measure of imbalance between elastic storage and viscous dissipation derived from magnetic resonance elastography (MRE). Twenty-eight patients (15 meningiomas, 13 glioblastomas) were analyzed using standardized 3 T MRE and tumor segmentation. Quantitative descriptors of instability topology—including skeleton length and branch-point densities, and radial persistence (radial-AUC)—were compared across WHO I, WHO II, and glioblastoma groups. Glioblastomas showed diffuse, branched instability fields with significantly higher skeleton and branch-point densities and lower radial-AUC compared with WHO I meningiomas, which exhibited compact, radially coherent patterns; WHO II meningiomas were intermediate. Group-average probability maps confirmed a transition from coherent to fragmented instability with increasing malignancy. These findings demonstrate that peritumoral mechanical topology reflects the degree of viscoelastic coupling at the tumor–brain interface. Instability mapping thereby extends conventional stiffness-based MRE metrics, offering a quantitative framework for assessing interface integrity and heterogeneity that may aid in elasticity-guided treatment strategies and biomechanical phenotyping of brain tumors.

**Statement of significance:** Intracranial tumors can disrupt the mechanical equilibrium of the brain, yet how these changes govern tissue deformation, invasion, and surgical behavior remains unclear. Here we introduce *mechanical instability mapping*—a framework that quantifies the spatial organization of viscoelastic imbalance around tumors using magnetic resonance elastography. Distinct topological patterns of instability differentiate benign from malignant lesions: meningiomas preserve coherent elastic coupling, whereas glioblastomas display fragmented, dissipative fields indicative of invasive, mechanically decoupled growth. These findings identify a new biomechanical signature of tumor–brain interaction and establish instability topology as a quantitative link between imaging, histopathology, and operative mechanics, with potential applications in elasticity-guided neurosurgery, biomechanical phenotyping, and MR-based histopathology.

## 1. Introduction

The brain is an exceptionally soft, viscoelastic organ whose structural integrity reflects a finely balanced interplay between its solid and fluid components. Foundational biophysics and biomechanics work shows that neural tissue exhibits pronounced nonlinearity, viscoelasticity, and poroelastic fluid–solid coupling across scales (from axons and extracellular matrix (ECM) to whole-brain response)^1–3^. When a tumor develops within this environment, it introduces marked mechanical heterogeneity—changing how stresses are transmitted, dissipated, and redistributed in the surrounding parenchyma. Such changes can influence tumor progression and the practical mechanics of surgical resection. Conceptually, a tumor and its microenvironment form a coupled viscoelastic system in which contrasts in elasticity, viscosity, and interfacial adhesion govern the deformation field at multiple scales^4,5^.

Tumor expansion perturbs local microarchitecture, compresses and remodels vasculature, and reorganizes the ECM. The ECM itself can stiffen or undergo architectural changes that alter stress propagation and mechanotransduction, shaping cell behavior and invasion ^4,6,7^. In gliomas, for example, abnormal ECM composition and mechanics have been linked to altered cell migration and diffuse infiltration^8^. At mesoscopic scales, heterogeneity in ECM viscoelasticity and fluid–matrix coupling lead to local contrasts in stress relaxation and energy dissipation. These contrasts can guide deformation along preferential paths, such that portions of the tumor–brain interface maintain coherent mechanical coupling while others decouple irregularly^5^.

Clinically, the consequences of this mechanical coupling are well recognized, as differences in stiffness and interface cohesion manifest directly in operative handling and surgical and treatment outcomes^9–13^. Low grade meningiomas are frequently firm and well circumscribed, presenting with a separable plane at the brain surface, whereas glioblastomas (GBMs) are more infiltrative and lack a distinct dissection plane. These observations align with differences in tumor–parenchyma mechanics reported in imaging and intraoperative series. Magnetic resonance elastography (MRE) studies, including those performed by this group, show that meningiomas are stiffer, while GBMs are softer and exhibit higher damping/viscous behavior than normal tissue^14–18^. Spatial variations in viscoelasticity are a feature of brain tumors, with mechanical heterogeneity often most pronounced near the tumor–parenchyma boundary^8,17,18^.

From a physical standpoint, the tumor–brain interface behaves as a boundary between two viscoelastic continua with contrasting mechanical properties. When this contrast exceeds a critical threshold, the interface may lose coherence and develop localized perturbations or slip bands, analogous to those observed in soft-matter systems under differential strain ^4,19–21^. Similar interfacial instabilities are well established in physics. The Saffman–Taylor instability in viscous flows describes the branching of an interface when a low-viscosity fluid displaces a more viscous one ^20,22,23^. In solids, Biot’s surface instability (1963) and subsequent theories of viscoelastic bifurcation demonstrate how competition between elastic storage and viscous dissipation gives rise to wrinkling, shear bands, and other localized deformation patterns^21,24^. In biological soft tissues, this same principle manifests through extracellular matrix remodeling and cytoskeletal reorganization under mechanical stress^8,25^.Collectively, these frameworks converge on a unifying principle: mechanical instability arises when the equilibrium between stored and dissipated energy is disrupted. Consequently, the morphology of high-instability regions observed in MRE maps may encode the topology of mechanical coupling and failure.

The aim of this study is to determine whether the spatial organization of peritumoral mechanical instability differs systematically between benign and malignant intracranial tumors, and whether these patterns offer a biomechanical explanation for distinct clinical behavior. We tested the following hypotheses:

i. WHO I meningiomas exhibit confined, symmetric peritumoral instability consistent with stable elastic coupling and a well-defined interface;
ii. WHO II meningiomas show locally expanded or irregular instability, indicating partial interface disruption;
iii. GBMs display diffuse, branched instability extending into surrounding parenchyma, consistent with infiltrative and more dissipative coupling.

## 2. Methods

### 2.1 Study design and participants

The study was conducted in accordance with the Declaration of Helsinki and approved by the Regional Committee on Health Research Ethics for the Region of Southern Denmark (IDs: S-20190105, S-20220055). All participants provided written informed consent prior to inclusion. Patients with suspected intracranial meningiomas scheduled for surgical resection at the Department of Neurosurgery, Odense University Hospital, Denmark, were consecutively recruited. For comparison, patients with newly diagnosed glioblastoma (GBM) were included from an independent prospective study applying the same MRE acquisition protocol^26^.

### 2.2 Image acquisition

MRE was performed on a 3 T system (Achieva, Philips Healthcare, The Netherlands) using a 16-channel head coil. The examination included a contrast-enhanced T1-weighted (T1W) gradient-echo sequence after intravenous gadobutrol (Gadovist 1 mmol/mL; Bayer, Germany) at 0.1 mL/kg. Shear waves at 60 Hz were generated using a pneumatic driver (Resoundant Inc., USA) positioned beneath the head, operated at approximately 20% of the manufacturer’s maximum amplitude.

Displacement fields were acquired using a single-shot spin-echo echo-planar MRE sequence (TR/TE = 4800/67 ms; field of view = 240 × 240 mm²; SENSE = 3; 48 slices; slice thickness = 3 mm; voxel = 3 mm³; 8 phase offsets).

Elastograms were reconstructed using a direct inversion algorithm (Yin et al., 2018) to yield maps of the storage modulus G′ (elasticity), loss modulus G″ (viscous damping), and the magnitude |G*|.

All maps were rigidly co-registered and resampled to the |G*| grid using 3D Slicer (v5.6) with visual verification^27^.

### 2.3 Tumor segmentation and region definition

For meningiomas, tumor segmentation was performed semi-automatically on contrast-enhanced T1W images in 3D Slicer. Boundaries were outlined manually (JSA, 3 years’ experience), refined by region-growing, and inspected manually. Ten tumor were resegmented after 12 weeks to assess intra-observer reliability.

For glioblastomas, ROIs were delineated independently by two observers—a research assistant (AIF, 3 years) and a neuroradiologist (FSH, 9 years)—including only contrast-enhancing solid tumor while excluding necrosis and cystic areas.

T1-based brain masks were generated using SynthStrip, refined to parenchyma masks, and eroded by 1 voxel for meningiomas to avoid partial-volume effects at the cortical surface^28^.

### 2.4 Definition of tumor rim and peritumoral shell

The rim region (r) was defined as the 1-voxel-thick layer immediately outside the tumor within the parenchyma.

The peritumoral shell (p) was defined as voxels located 0–6 mm from the tumor boundary, excluding voxels within 2 mm of the skull. Distances were computed in physical mm using an anisotropy-aware Euclidean distance transform.

These definitions were applied identically to meningiomas and GBMs.

### 2.5 Mechanical instability field

Mechanical instability was modeled as a voxel-wise scalar field I(x) describing the local imbalance between elastic storage and viscous dissipation relative to the rim reference.

Within the rim (r), the median loss modulus G″₍r, med₎ and median damping ratio (tan δ)₍r, med₎ were computed after all zero or negative value voxels were excluded from the ROIs.

For each voxel x in the peritumoral shell (p), instability was defined as:

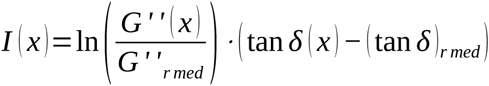

where G″(x) and tan δ(x) are the local loss modulus and damping ratio.

The logarithmic term quantifies the viscous contrast, and the phase-lag term captures local differences in dissipation.

Resulting maps were masked to parenchyma, saved as NIfTI volumes, and visualized alongside G′, G″, and T1W images with overlaid contours.

### 2.6 Instability-derived metrics

From each subject’s peritumoral domain, a series of quantitative descriptors were derived to summarize the magnitude, spatial extent, and topology of mechanical instability. The median and 95th percentile of the instability field *I* (*x*) were calculated, together with the fraction of voxels exceeding the thresholds I > 0.02 and I > 0.05. The cutoff for defining instability (I > 0.02) was chosen from the pooled histogram of all cases at the point where background fluctuations gave way to coherent peritumoral structures. The entropy of the histogram (32 bins, range 0–0.4) quantified mechanical heterogeneity within the peritumoral region.

To characterize the radial organization of instability, profiles *I* (*r*) were computed in 2-mm concentric shells extending up to 12 mm from the tumor boundary only including parenchyma. The area under this curve (AUC) represented the persistence of mechanical perturbation with distance. The integrated survival function *S* (*t*)=*P* (*I* >*t*)up to I = 0.30 provided a complementary measure of the overall extent of instability, reported as the Tail-AUC.

Morphological properties of the instability field were analyzed on the axial slice containing the maximal tumor area. Thresholded maps (I > 0.02) were used to compute the *isoperimetric ratio* (*IPR*)=4 *π A* / *P*^2^, reflecting boundary irregularity, and convexity, defined as the ratio between the region area and the area of its convex hull (*A* / *A_convex_*).

To capture the internal topology of mechanically unstable regions, each thresholded map was converted into a binary mask and skeletonized using an 8-connected morphological algorithm implemented in *scikit-image*. The resulting one-voxel-wide skeleton preserved the medial axis of each unstable component. Skeleton length density was computed as the total number of skeleton voxels divided by the number of voxels in the unstable region, representing the normalized total length of unstable filaments per unit area. Branch-point density was defined as the proportion of skeleton voxels with three or more neighboring skeleton voxels (in 8-connectivity), quantifying the degree of branching and fragmentation of the instability network.

Together, these parameters describe the tortuosity, fragmentation, and spatial persistence of the peritumoral mechanical field. Per-case values were subsequently aggregated by diagnostic group (WHO I, WHO II, GBM) for statistical comparison.

### 2.7 Group-average instability and skeleton maps

To visualize group-level patterns, each subject’s instability map *I(x)* was cropped to a 120 × 120 mm physical field centered on the axial slice containing the maximal tumor cross-section and resampled to a uniform 256 × 256 grid in physical space. Voxels exceeding the instability threshold (I > 0.02) were binarized, skeletonized using 8-connectivity, and branch points identified where three or more skeleton segments converged.

For each group g ∈ {WHO I, WHO II, GBM}, occupancy O_i_, skeleton S_i_, and branch B_i_ maps were averaged voxelwise over n_g cases to yield group-probability maps:

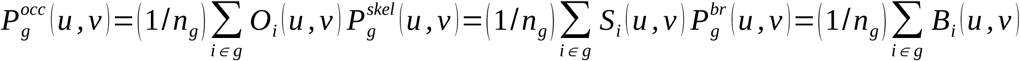

Group maps were displayed with fixed ranges (0–1.0, 0–0.8, 0–0.6) for direct comparison.

The resulting figure illustrates the probability and geometric complexity of unstable regions in WHO I, WHO II, and GBM tumors.

### 2.8 Robustness analyses

To ensure that group differences in instability topology were not driven by arbitrary parameter choices, we performed a series of robustness checks. First, we varied the instability cutoff over a plausible range (I > 0.015, 0.020, 0.025, 0.030). For each threshold, per-case topology metrics—radial persistence (radial-AUC), skeleton length density, and branch density—were recomputed within the 0–6 mm peritumoral shell. Second, to assess sensitivity to pixel-scale irregularities, skeletonization was repeated after a light binary morphology opening (1.5 mm in-plane).

Third, we quantified the fraction of negative instability values (pₙₑg) per case to indicate how much of the peritumoral field behaved elastically (mechanically stable) versus dissipatively. Fourth, to examine potential size bias, we evaluated correlations between topology metrics (e.g., radial-AUC) and tumor size proxies such as shell voxel count or tumor area.

Finally, to verify that spatial topology was not confounded by tumor scale, we generated *radius-normalized group probability maps* in which each tumor was rescaled to a common effective radius (R* = 20 mm) prior to averaging.

To relate local topology to bulk mechanical contrast, we computed rim–shell differences in storage modulus and damping *ΔG’* =*G’* _rim_ *−G’*_shell_ *; Δ* tan *δ* =(tan *δ*)_rim_ *−* (tan *δ*)_shell_, merged these with per-case topology metrics, and evaluated associations using Spearman correlation.

Per-case metrics were summarized by their median across thresholds to avoid single-cutoff dependence.

### 2.9 Statistical analyses

Computation and statistical analyses were performed in Python (v3.11) using pandas, scipy.stats, and matplotlib.

Distributions of continuous variables were assessed visually by histograms and Q–Q plots. Because most parameters showed non-normal distributions, non-parametric tests were applied throughout.

For each instability-derived metric (tail-AUC, proportion above 0.02 and 0.05, entropy, isoperimetric ratio, convexity, skeleton branch-point density, skeleton length density, and radial AUC), group differences were evaluated using the Kruskal–Wallis test.

If the overall test reached significance (p < 0.05), pairwise comparisons between WHO I, WHO II, and GBM were conducted with two-sided Mann–Whitney U tests.

Multiple testing was controlled using the Benjamini–Hochberg false discovery rate (FDR) procedure applied separately across all pairwise contrasts and metrics.

The adjusted q-value threshold for significance was q < 0.05.

Effect sizes for pairwise comparisons were expressed as Cliff’s delta (δ), interpreted as negligible (|δ| < 0.147), small (0.147–0.33), medium (0.33–0.474), or large (>0.474).

The Hodges–Lehmann estimator (HL) of the median difference was additionally reported to indicate the magnitude and direction of the shift between groups.

## 3. Results

### 3.1 Cohort

Twenty-eight tumors were analyzed: fifteen meningiomas (10 WHO grade I, 5 WHO grade II) and thirteen GBMs (**Table 1**). All datasets met predefined image quality and segmentation criteria, and all cases were successfully processed through the voxelwise instability pipeline. **Figure 1** display the analytical flow.

**Figure 1.**
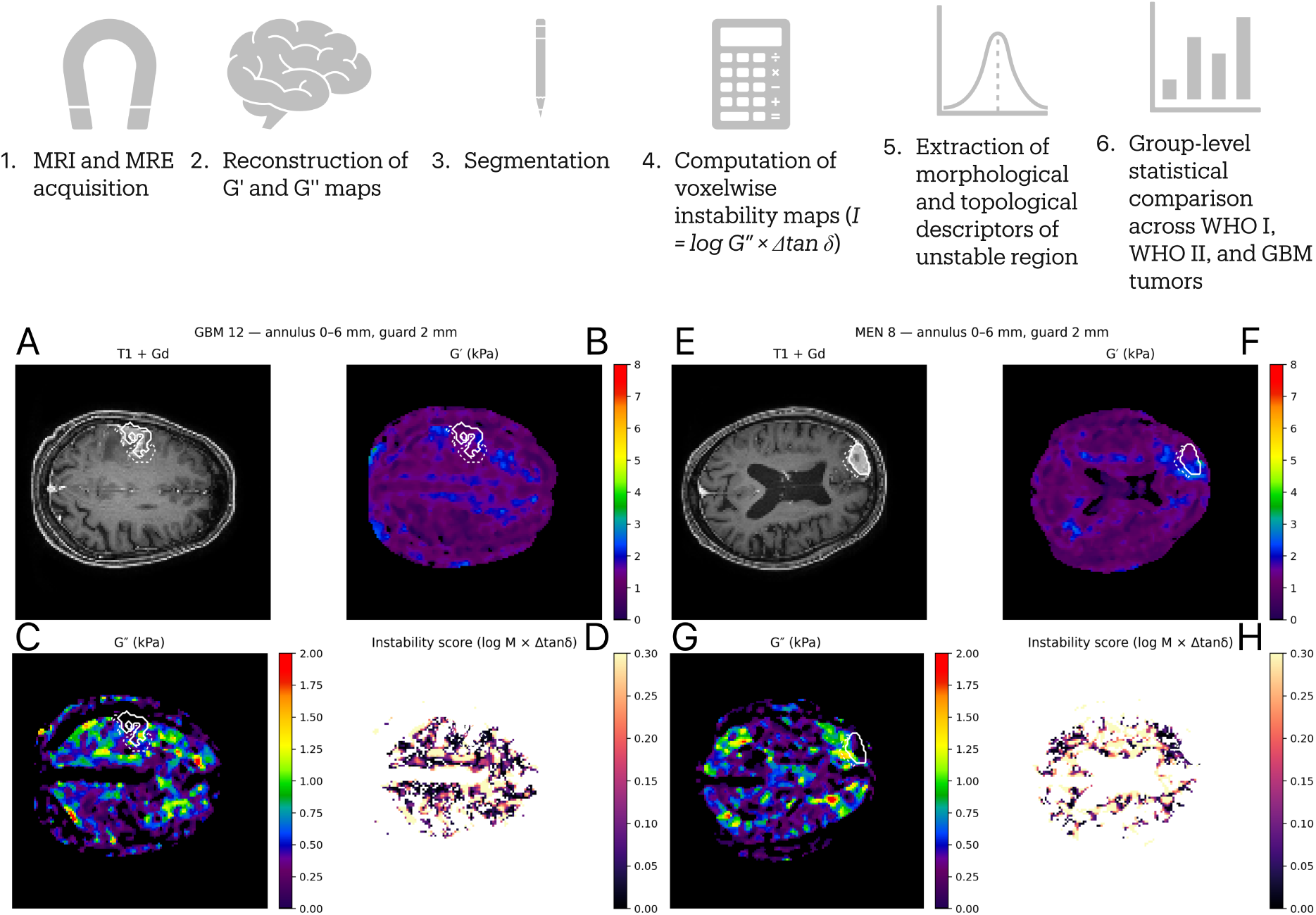
**Overview of the analytical workflow and representative examples of mechanical instability mapping in glioblastoma and meningioma**. The top panel summarizes the analytical pipeline, beginning with MRI and magnetic resonance elastography (MRE) acquisition, reconstruction of viscoelastic maps of storage (G′) and loss (G″) moduli, and tumor segmentation on contrast-enhanced T1-weighted images. Voxelwise mechanical instability was computed within the peritumoral annulus (0–6 mm, guard 2 mm) as the instability index (I), followed by extraction of morphological and topological descriptors for group-level comparison across WHO I, WHO II, and glioblastoma (GBM) cohorts. The lower panels show representative examples from a GBM (A–D) and a WHO I meningioma (E–H), displaying the contrast-enhanced T1-weighted image, G′ and G″ maps, and the derived instability score. White contours delineate the tumor and analysis shell.

**Table 1.**
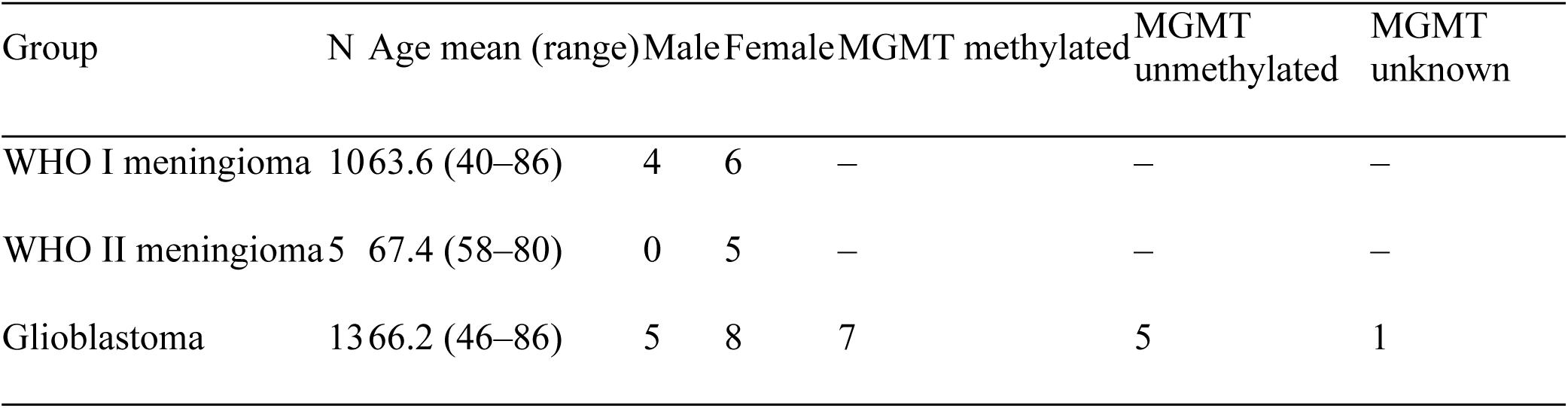
Cohort characteristics.

### 3.2 Groupwise distributions of instability metrics

Descriptive metrics summarizing the topology of peritumoral instability are reported in **Table 2**. Groupwise differences were observed across diagnostic categories. Median skeleton branch-point density increased progressively from 0.015 in WHO I meningiomas to 0.029 in WHO II and 0.151 in GBMs, indicating an increasing number of local bifurcations within the unstable region. Median skeleton length density followed a similar pattern, increasing from 0.126 (WHO I) to 0.324 (WHO II) and 0.326 (GBM), suggesting a greater cumulative length of unstable structures per unit area in higher-grade lesions. Conversely, radial-AUC, which quantifies the persistence of instability with distance from the tumor margin, was highest in WHO I meningiomas (1.193), lower in GBM (0.714), and lowest in WHO II (0.546).No substantial group differences were noted for convexity or isoperimetric ratio, with median convexity values ranging from 0.50 to 0.54 and IPR values from 8.7 to 18.1 across groups.

**Table 2.**
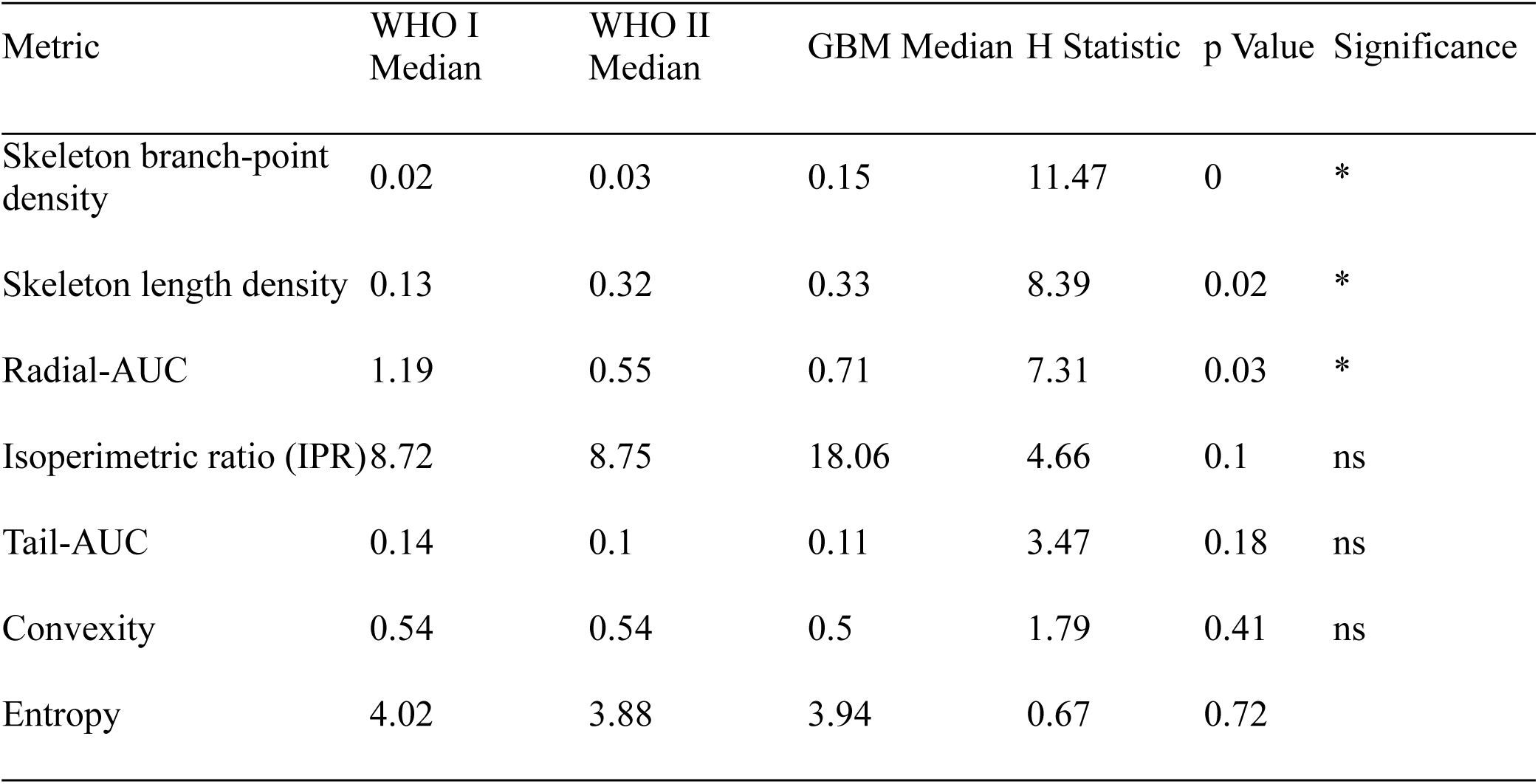
Groupwise medians and Kruskal–Wallis test results for all instability-derived metrics.

### 3.3 Kruskal–Wallis group effects

Nonparametric Kruskal–Wallis tests revealed statistically significant overall group effects for skeleton branch-point density (H = 11.47, p = 0.0032), skeleton length density (H = 8.39, p = 0.0151), and radial-AUC (H = 7.31, p = 0.0258). No significant group effects were observed for entropy (H = 0.67, p = 0.716), tail-AUC (H = 3.47, p = 0.177), isoperimetric ratio (H = 4.66, p = 0.097), or convexity (H = 1.79, p = 0.408). These results are summarized in **Table 2** and the distribution is visualized in **Figure 2**.

**Figure 2.**
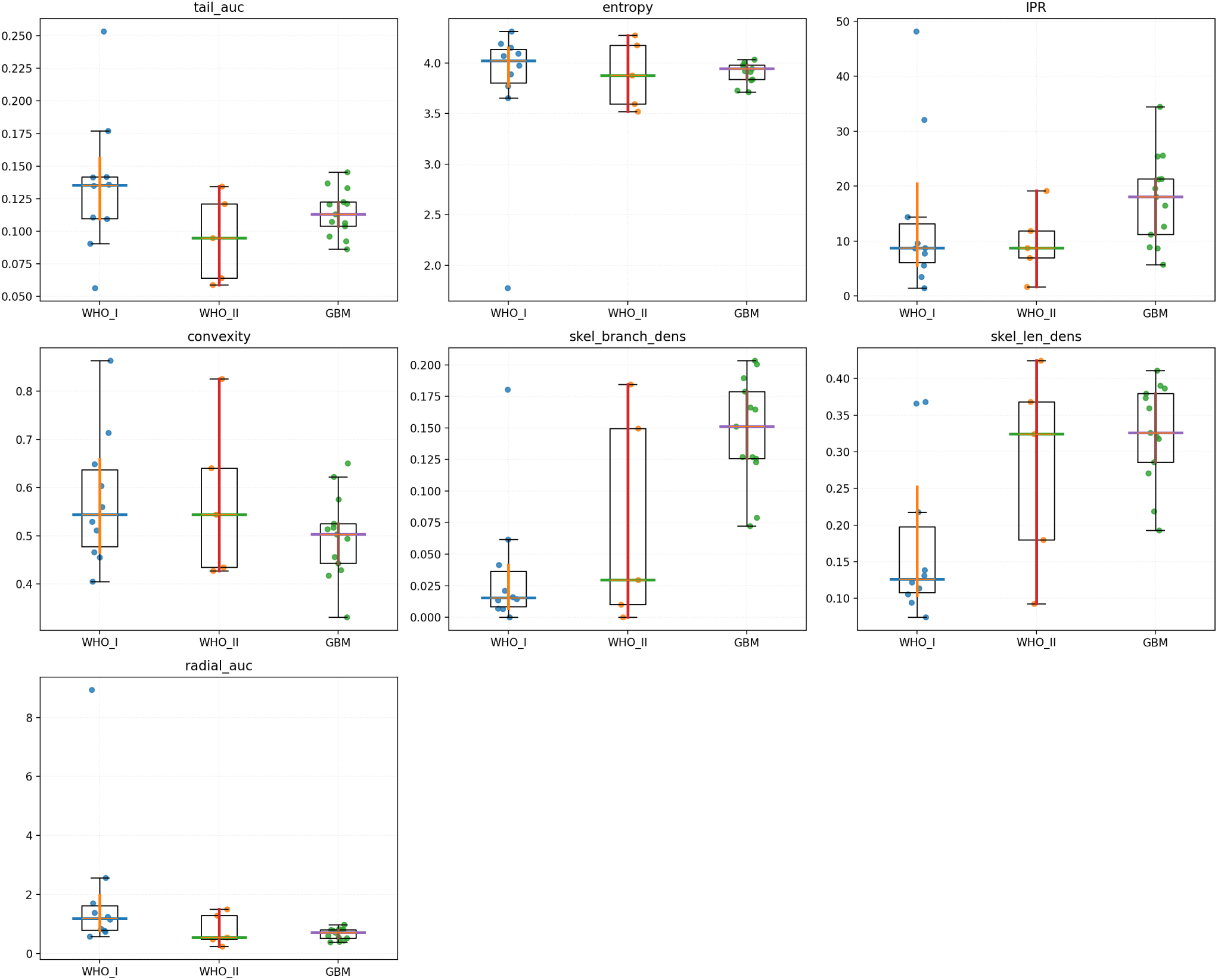
Groupwise distributions of instability-derived metrics. Box-and-swarm plots show metrics summarizing peritumoral mechanical instability in WHO I meningiomas (blue), WHO II meningiomas (orange), and glioblastomas (green). Horizontal bars indicate medians with bootstrap 95% CIs. Significant overall group effects (Kruskal–Wallis) were observed for skeleton branch-point density, skeleton length density, and radial-AUC (all p<0.05); other metrics were not significant. Pairwise Mann–Whitney tests with BH-FDR correction identified WHO I vs GBM differences for radial-AUC (WHO I > GBM), branch-point density (WHO I < GBM), and skeleton length density (WHO I < GBM).

Pairwise Mann–Whitney U tests followed by Benjamini–Hochberg false discovery rate (FDR) correction (q < 0.05) identified three significant contrasts:

1. Radial-AUC: WHO I > GBM (U = 109, p = 0.007, q = 0.049, Cliff’s δ = –0.68, Hodges–Lehmann shift = +0.46).

WHO I meningiomas exhibited greater radial persistence of instability relative to GBM.

2. Skeleton branch-point density: WHO I < GBM (U = 10, p = 7.3 × 10⁻⁴, q = 0.015, δ = +0.85, HL = –0.12).

GBMs showed a substantially higher density of branch points within unstable regions.

3. Skeleton length density: WHO I < GBM (U = 17, p = 0.0032, q = 0.034, δ = +0.74, HL = – 0.18).

GBMs presented longer cumulative skeleton structures per area compared to WHO I lesions.

All Mann–Whitney U statistics, adjusted q-values, effect sizes, and Hodges–Lehmann median shifts are reported in **Table 3**.

**Table 3.**
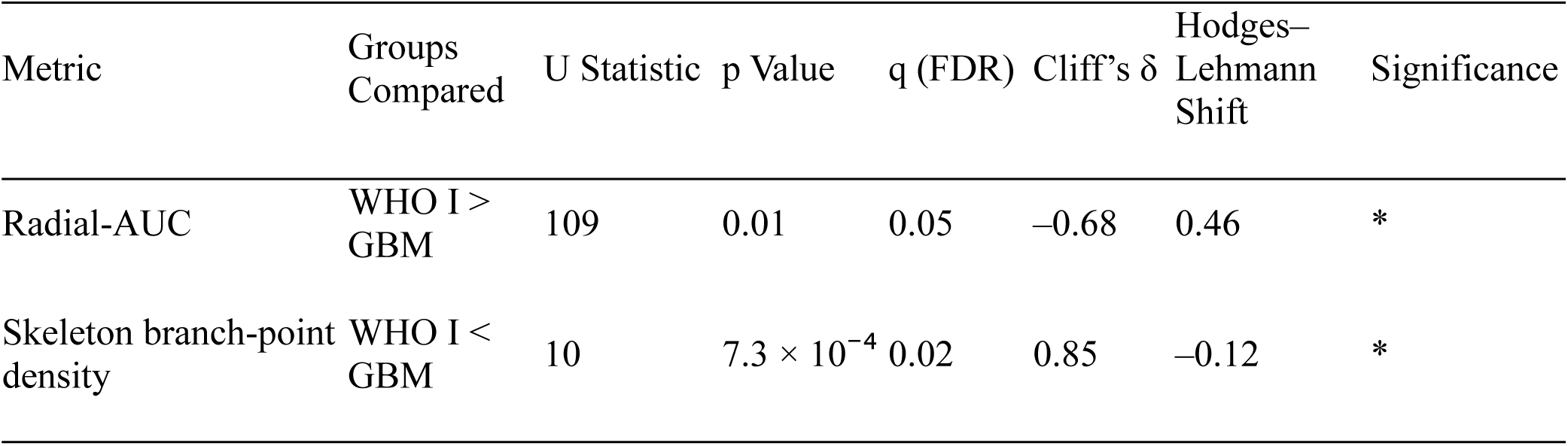
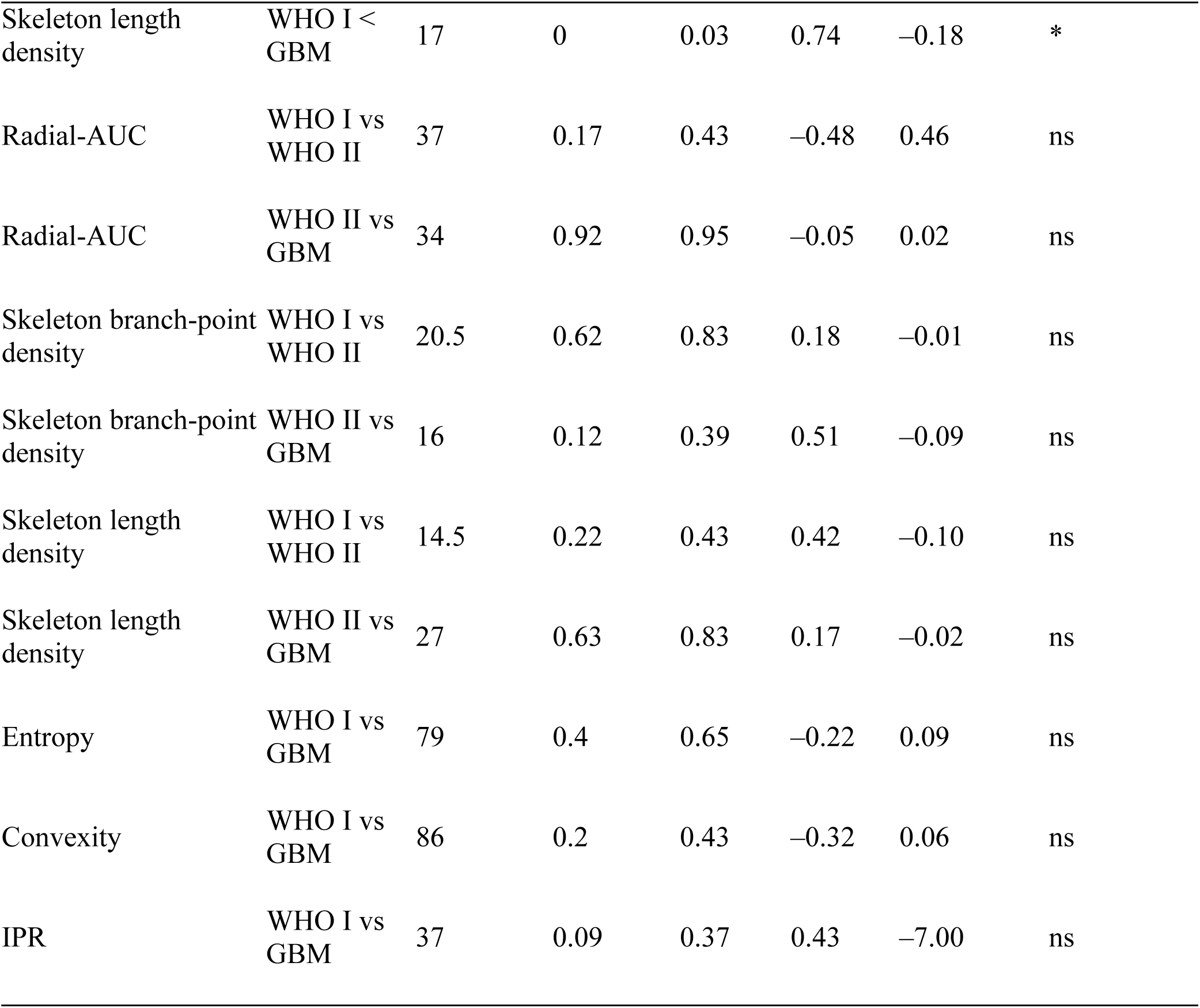
Pairwise Mann–Whitney U comparisons with Benjamini–Hochberg FDR correction.

| Metric | Groups Compared | U Statistic | p Value | q (FDR) | Cliff's $\delta$ | Hodges–Lehmann Shift | Significance |
| --- | --- | --- | --- | --- | --- | --- | --- |
| Radial-AUC | WHO I > GBM | 109 | 0.01 | 0.05 | –0.68 | 0.46 | * |
| Skeleton branch-point density | WHO I < GBM | 10 | $7.3 \times 10^{-4}$ | 0.02 | 0.85 | –0.12 | * |
| Skeleton length density | WHO I < GBM | 17 | 0 | 0.03 | 0.74 | −0.18 | * |
| Radial-AUC | WHO I vs WHO II | 37 | 0.17 | 0.43 | −0.48 | 0.46 | ns |
| Radial-AUC | WHO II vs GBM | 34 | 0.92 | 0.95 | −0.05 | 0.02 | ns |
| Skeleton branch-point density | WHO I vs WHO II | 20.5 | 0.62 | 0.83 | 0.18 | −0.01 | ns |
| Skeleton branch-point density | WHO II vs GBM | 16 | 0.12 | 0.39 | 0.51 | −0.09 | ns |
| Skeleton length density | WHO I vs WHO II | 14.5 | 0.22 | 0.43 | 0.42 | −0.10 | ns |
| Skeleton length density | WHO II vs GBM | 27 | 0.63 | 0.83 | 0.17 | −0.02 | ns |
| Entropy | WHO I vs GBM | 79 | 0.4 | 0.65 | −0.22 | 0.09 | ns |
| Convexity | WHO I vs GBM | 86 | 0.2 | 0.43 | −0.32 | 0.06 | ns |
| IPR | WHO I vs GBM | 37 | 0.09 | 0.37 | 0.43 | −7.00 | ns |

### 3.4 Group-average spatial probability maps

Group-average instability and skeleton probability maps are shown in **Figure 3**.

**Figure 3.**
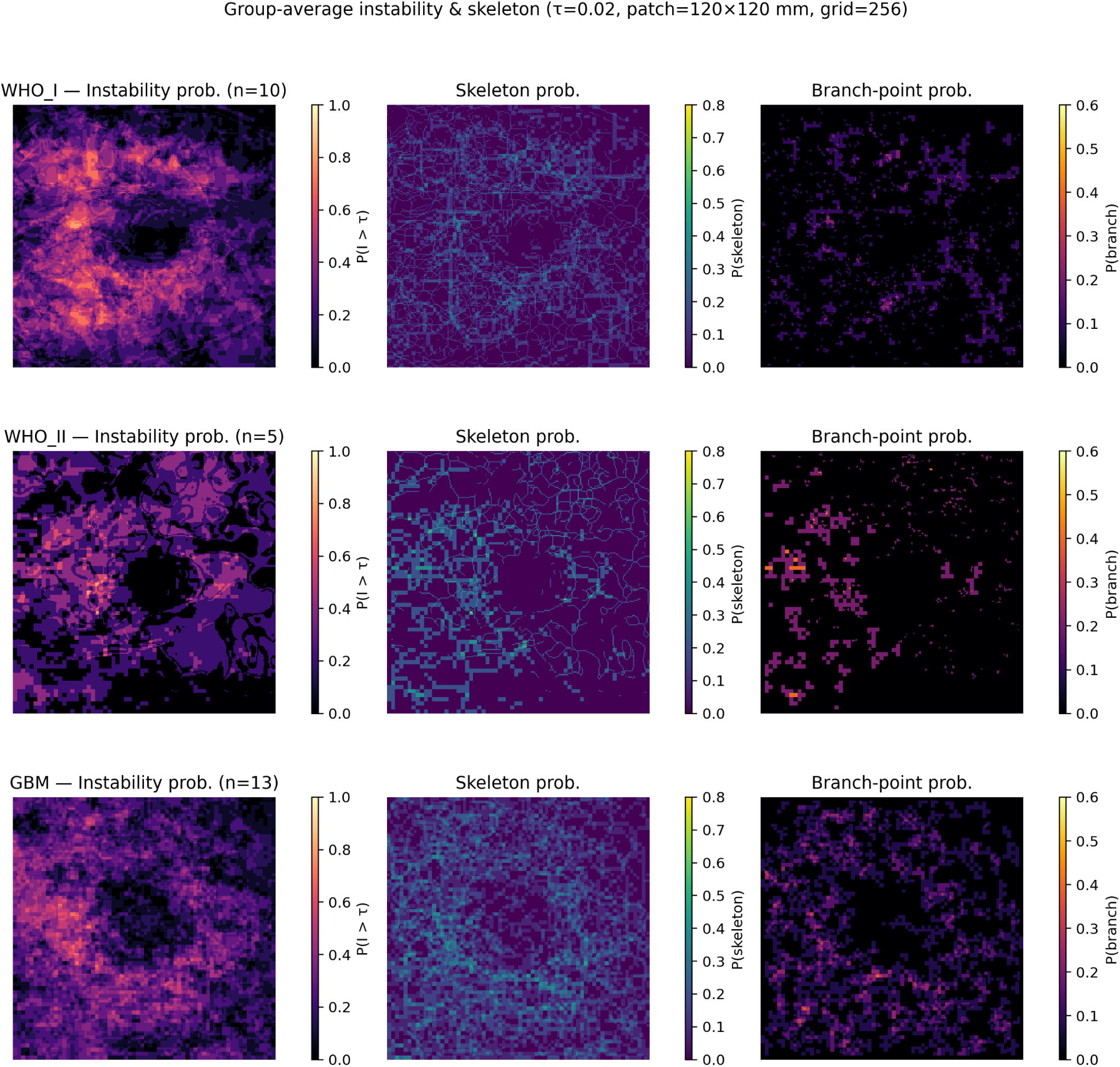
Group-average peritumoral instability and skeleton maps in WHO I, WHO II, and glioblastoma (GBM) Voxelwise instability maps were aligned to a common 120 × 120 mm physical patch centered on the slice of maximal tumor area and resampled to a 256 × 256 grid. Voxels exceeding the instability threshold (I > 0.02) were binarized and skeletonized to identify filamentous structures, with branch points defined where three or more filaments intersected. Group-average probability maps show the spatial frequency of instability (left), skeleton (middle), and branch-point occurrence (right) across subjects within each diagnostic group. Distances are expressed in millimeters, enabling direct spatial comparison across cases. The central dark region marks the excluded tumor core, which appears larger in GBM due to greater mean tumor size. Because maps were binarized before averaging, all subjects contribute equally. By counting only voxels within valid tissue regions, each location reflects the true probability of instability within brain tissue, rather than a size-weighted blend of valid and background areas.

Each heatmap represents the voxelwise average of binarized instability (I > 0.02), skeleton, and branch-point maps across all subjects within each diagnostic group, resampled to a common 120 × 120 mm physical grid.

Instability occupancy was most spatially compact in WHO I meningiomas, showed irregular expansion in WHO II, and appeared diffuse and branched in GBMs.

Skeleton and branch-point probability increased progressively with malignancy, consistent with the quantitative metrics summarized above.

### 3.5 Robustness

Varying the instability threshold from 0.015 to 0.030 did not alter group ordering or qualitative patterns. GBMs consistently showed more diffuse peritumoral fields (lower radial-AUC than WHO I meningiomas), while WHO II cases remained intermediate. Effect-size differences across thresholds were small (< 10%), indicating that results were not driven by a specific cutoff.

Applying a light morphology opening before skeletonization changed median skeleton metrics by < 8% and preserved group rankings, showing that filament and branch descriptors were not artifacts of pixel-scale noise.

Negative I(x) voxels accounted for 18 ± 7 % across cases (WHO I ≈ 21 %, GBM ≈ 15 %) confirming that peritumoral behavior is largely dissipative. These values indicate more localized stability in meningiomas and a broader dissipative field in GBMs. Median radial-AUC increased slightly with threshold across all groups but maintained the same ordering (GBM < WHO II < WHO I). Typical medians were ∼2.2–2.5 for GBM, ∼2.4–2.7 for WHO II, and ∼2.9–3.1 for WHO I. Radial-AUC showed only weak associations with tumor size proxies (|ρ| < 0.25), confirming that group averages in physical space were not dominated by larger tumors.

Radius-normalized probability maps reproduced the same spatial organization observed in physical coordinates—compact and symmetric for WHO I, heterogeneous for WHO II, and diffuse and branched for GBM—demonstrating that the observed topology differences are intrinsic rather than scale-dependent (**S1**)

Stiffness contrast (ΔG′) correlated inversely with radial-AUC (ρ = –0.46, p = 0.014), indicating that higher stiffness contrast is associated with a more compact, rim-confined instability field. Δtan δ showed no significant associations, and neither contrast predicted skeleton or branch-point densities (**S2**). N-maps are available in the supplementary material (**S3**) and visualize how many samples contributed to each voxel by group..

Together, these analyses confirm that group differences in instability topology—compact, radially coherent fields in WHO I meningiomas; broader, irregular fields in WHO II; and diffuse, fragmented fields in GBM—are robust to thresholding, morphological filtering, and size normalization, reflecting genuine biomechanical organization rather than parameter choice or geometric bias.

## 4. Discussion

This study establishes a quantitative framework for characterizing peritumoral mechanical instability from MRE and demonstrates distinct spatial organizations of viscoelastic imbalance across intracranial tumor types. By modeling instability as a voxelwise divergence between elastic storage and viscous dissipation, we show that benign and malignant tumors exhibit fundamentally different modes of mechanical coupling to the surrounding brain.

In WHO I meningiomas, instability fields were spatially compact, smooth, and radially coherent, with high radial-AUC and low skeleton densities. In contrast, GBMs displayed diffuse, branched instability networks with markedly higher skeleton length and branch-point densities and a rapid radial decay. WHO II meningiomas occupied an intermediate position between these extremes. These findings indicate that tumors with higher invasive potential exhibit reduced mechanical coherence and greater viscoelastic dissipation at the brain interface.

### 4.1 Mechanical coupling and interface coherence

The tumor–brain interface represents a boundary between two viscoelastic continua. When elastic and viscous stresses remain balanced, the interface deforms coherently; when this balance is disrupted, energy is dissipated through localized deformation or slip ^21,23,24^. The confined instability patterns in meningiomas suggest that elastic storage dominates near the interface, preserving structural coherence and a predictable dissection plane. This elastic dominance corresponds to a mechanically stable configuration, consistent with high stiffness and low damping in MRE studies and with the absence of Saffman–Taylor-type instabilities. In contrast, glioblastomas, which exhibit lower stiffness and reduced viscous storage, fulfill the conditions for viscous fingering and display diffuse, branched instability fields consistent with interface decoupling and infiltrative behavior ^29^. Conversely, the fragmented instability topology in GBM aligns with a more viscous, energy-dissipative microenvironment where coupling fails locally, analogous to branching seen in soft-matter systems under differential strain ^4,20,23^. The mechanical transition observed here—from coherent to branched instability—may therefore represent a physical signature of viscoelastic decoupling during malignant invasion.

### 4.2 Topological metrics as descriptors of viscoelastic heterogeneity

Skeleton length and branch-point densities provided compact descriptors of the topology of mechanical instability. These parameters quantify the complexity of the unstable network rather than its amplitude, thereby capturing fragmentation of mechanical coherence. Their progressive increase from WHO I to GBM parallels histological observations of irregular ECM architecture and variable adhesion at malignant interfaces^8,25^. Radial-AUC, conversely, describes the spatial persistence of instability away from the tumor margin, indicating the extent to which mechanical perturbations penetrate adjacent brain. Together, these metrics define complementary axes of interface mechanics: *coherence* (radial-AUC) and *fragmentation* (skeleton topology). Their combination offers a quantitative language for describing tumor–parenchyma coupling, analogous to stiffness and damping parameters but with spatial context.

### 4.3 Clinical and translational significance

Understanding how tumors mechanically interact with surrounding tissue has direct relevance for neurosurgical tumor resection and for emerging clinical elasticity-guided strategies^10,29^. Compact, coherent instability fields—as in low-grade meningiomas—imply predictable deformation behavior and a stable cleavage plane, whereas diffuse and branched patterns—as in GBM—suggest heterogeneous spread and potential for irregular motion or residual infiltration. Quantitative instability mapping could therefore complement conventional stiffness measures in preoperative assessment, predicting micro-invasion, recurrence etc.

Local variations in the instability field may also reflect boundary conditions imposed by nearby anatomical structures such as ventricles or major white-matter tracts. These local constraints could influence how tumors deform or invade surrounding tissue, effectively channeling growth along paths of least mechanical resistance.

Beyond resection planning, instability topology provides a framework for linking in vivo mechanics to extracellular matrix remodeling and invasive potential^6,8,25^. When combined with diffusion imaging, radiomics, or molecular data, these maps could improve non-invasive tumor phenotyping and treatment stratification. Longitudinal monitoring of instability patterns may further enable early detection of biomechanical changes that precede radiographic progression or therapeutic response, and be used to predict the growth pattern^21,23,24^. Spatial patterns of instability might help explain where tumors tend to extend or recur after treatment. Such integrative, elasticity-informed approaches could ultimately refine surgical strategy, guide therapy, and support the development of mechanically targeted interventions in neuro-oncology.

### 4.4 Future directions

Future work should integrate instability mapping with histological, proteomic, and rheometric data to elucidate the biochemical substrates of mechanical fragmentation. Coupling instability fields with dynamic MRE at multiple drive frequencies may also reveal frequency-dependent bifurcation behavior and fluid–solid coupling in vivo. Longitudinal monitoring could determine whether changes in instability topology accompany treatment response or recurrence, potentially establishing a biomechanical biomarker of therapeutic effect and prediction of the growth pattern.

### 4.5 Methodological considerations and limitations

The instability index I(x) simplifies complex viscoelastic behavior into a scalar measure of local imbalance. While this abstraction omits directional and anisotropic effects, it enables reproducible voxelwise comparison across patients. The spatial domain was chosen to capture the immediate mechanical transition zone, though more extensive fields could reveal longer-range poroelastic interactions. Patch-based alignment was performed in 2D to preserve spatial interpretability, but future 3D registration could better account for tumor orientation. The moderate sample size limits detection of subtler differences, particularly between WHO I and II lesions, and validation in large cohorts is warranted.

## 5. Conclusions

Peritumoral mechanical instability mapping reveals systematic differences in the spatial organization of viscoelastic imbalance between WHO I- and WHO II meningiomas, and GBMs. Low-grade meningiomas exhibit confined, coherent, and radially ordered instability consistent with stable elastic coupling, whereas GBMs show diffuse, branched, and fragmented patterns indicative of dissipative decoupling. These observations link macroscopic mechanical topology to tumor biology and operative and invasive behavior, suggesting that instability mapping may serve as a quantitative tool for assessing tumor–brain interface integrity and guiding elasticity-based strategies.

## Data Availability

All data produced in the present study are available upon reasonable request to the authors

## Acknowledgments

This work was supported by the Danish Cancer Society (grants R326-A19155 and R352-A20745). Additional support was provided by the National Institutes of Health (NIH) through grants R37-EB001981 and R01-NS113760. We thank Odense University Hospital, the staff of the Department of Neurosurgery and the Department of Radiology, for their collaboration and technical assistance. Finally, we wish to acknowledge the contribution of Richard L. Ehman, M.D. and John Huston, III, M.D for facilitating collaboration between the contributing institutions.

## Disclosure

During the preparation of this work, the authors used ChatGPT-5.0 (OpenAI) to assist with language refinement and title/abstract drafting, and GitHub Copilot (GitHub) to assist with boilerplate Python code generation for data processing scripts. The authors reviewed and edited all content and take full responsibility for the integrity and accuracy of the final manuscript.

---

**Supplementary Figure S1.**
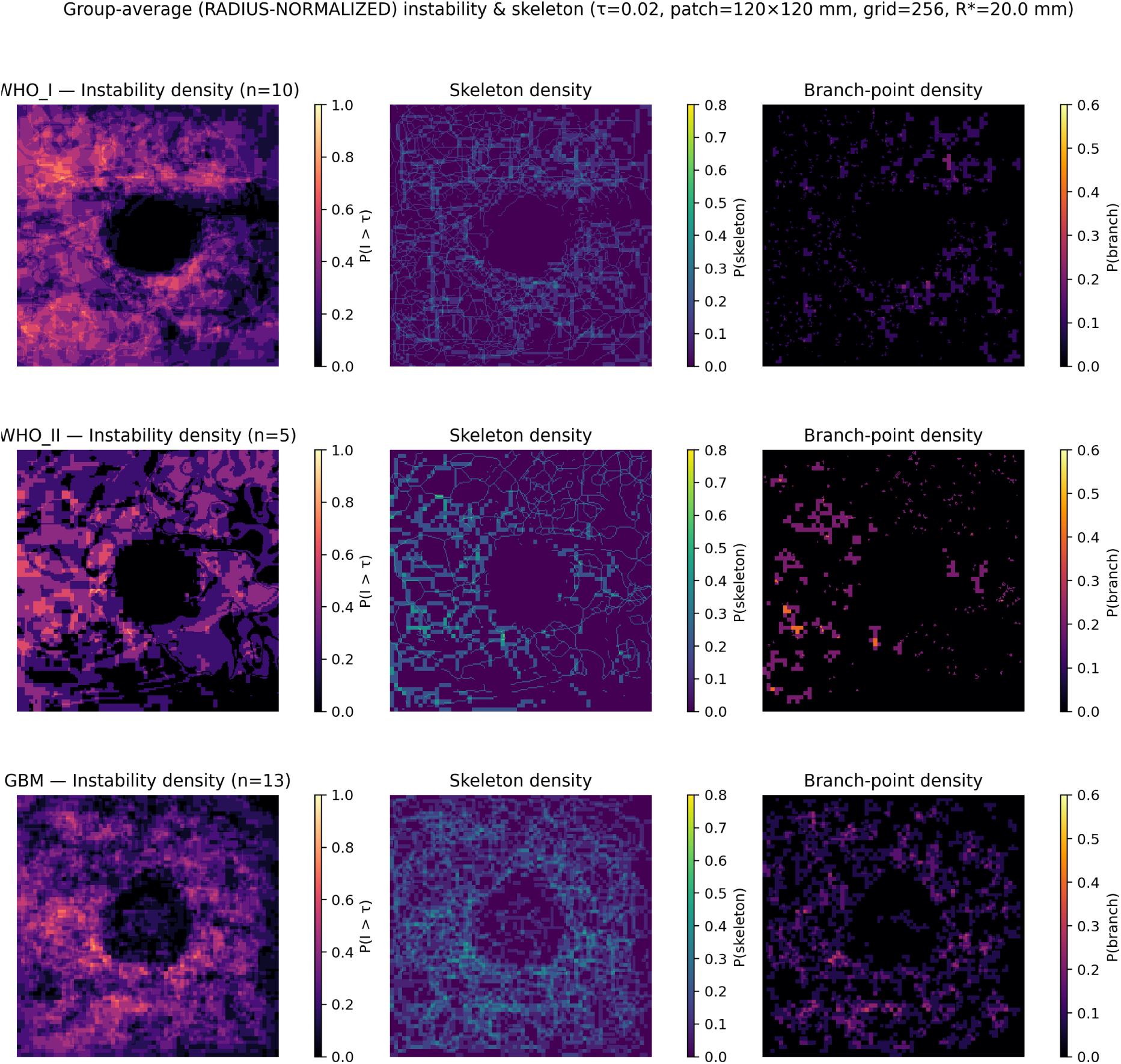
Radius-normalized group maps of instability topology. Group-average maps showing the spatial distribution of peritumoral instability after radius normalization to a common effective tumor radius (R* = 20 mm).Each row corresponds to a diagnostic group: WHO I, WHO II, and GBM. Columns show (left) instability density (probability of I > τ = 0.02), (middle) skeleton density (probability of filament presence), and (right) branch-point density (probability of bifurcations). Compact, radially symmetric instability fields characterize WHO I meningiomas, whereas higher-grade lesions show broader and more fragmented patterns.

**Supplementary Figure S2.**
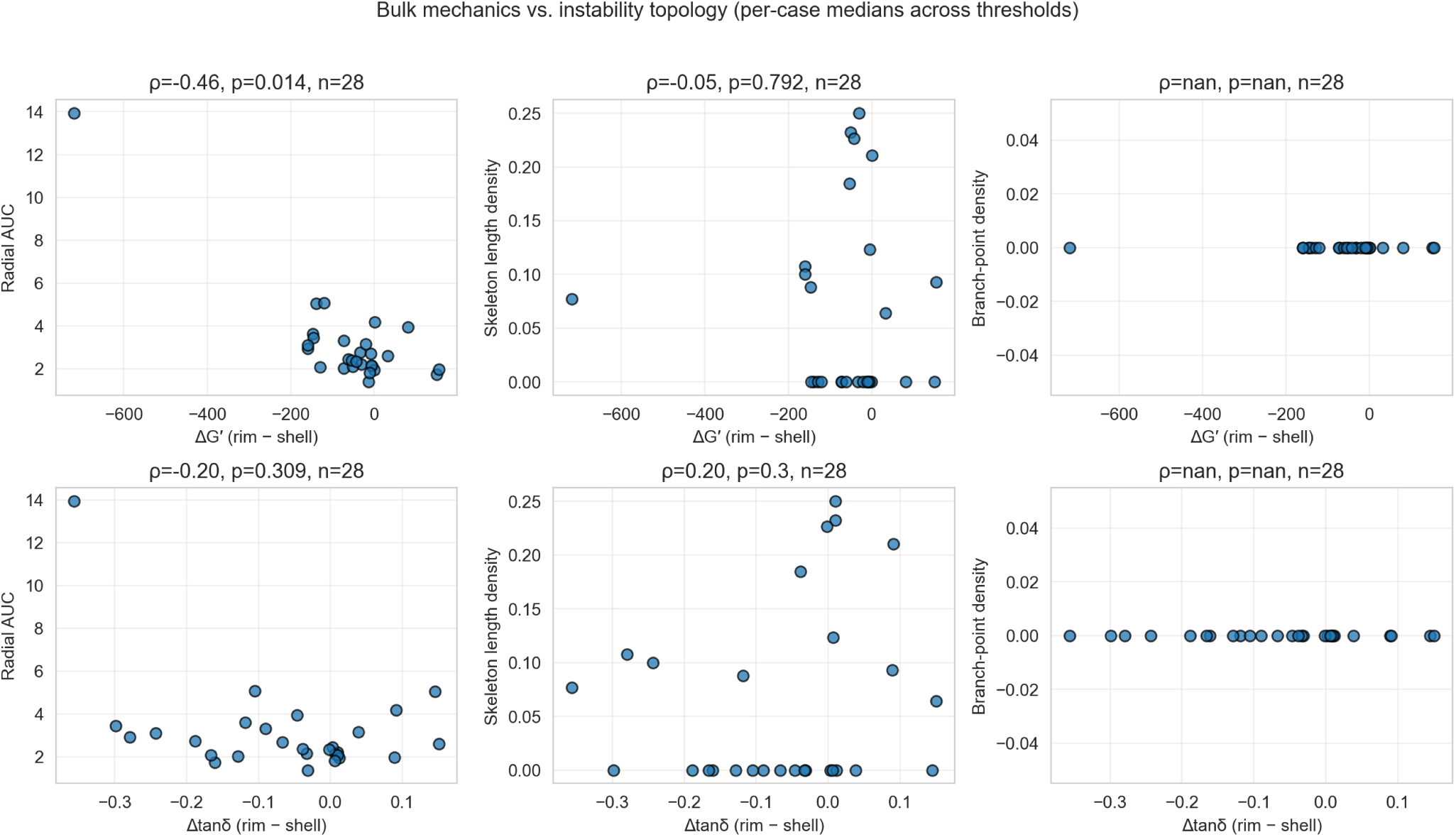
Relationship between bulk mechanics and instability topology. Scatter-matrix showing correlations between rim–shell mechanical contrasts—ΔG′ = G′₍rim₎ – G′₍shell₎ and Δtan δ = tan δ₍rim₎ – tan δ₍shell₎—and topology metrics (radial-AUC, skeleton length density, branch-point density). Each panel reports the Spearman correlation coefficient (ρ), p-value, and number of cases (n = 28). A significant inverse relationship was observed between ΔG′ and radial-AUC (ρ = –0.46, p = 0.014), indicating that higher stiffness contrast corresponds to more compact instability fields. No other associations reached significance.

**Supplementary Figure S3.**
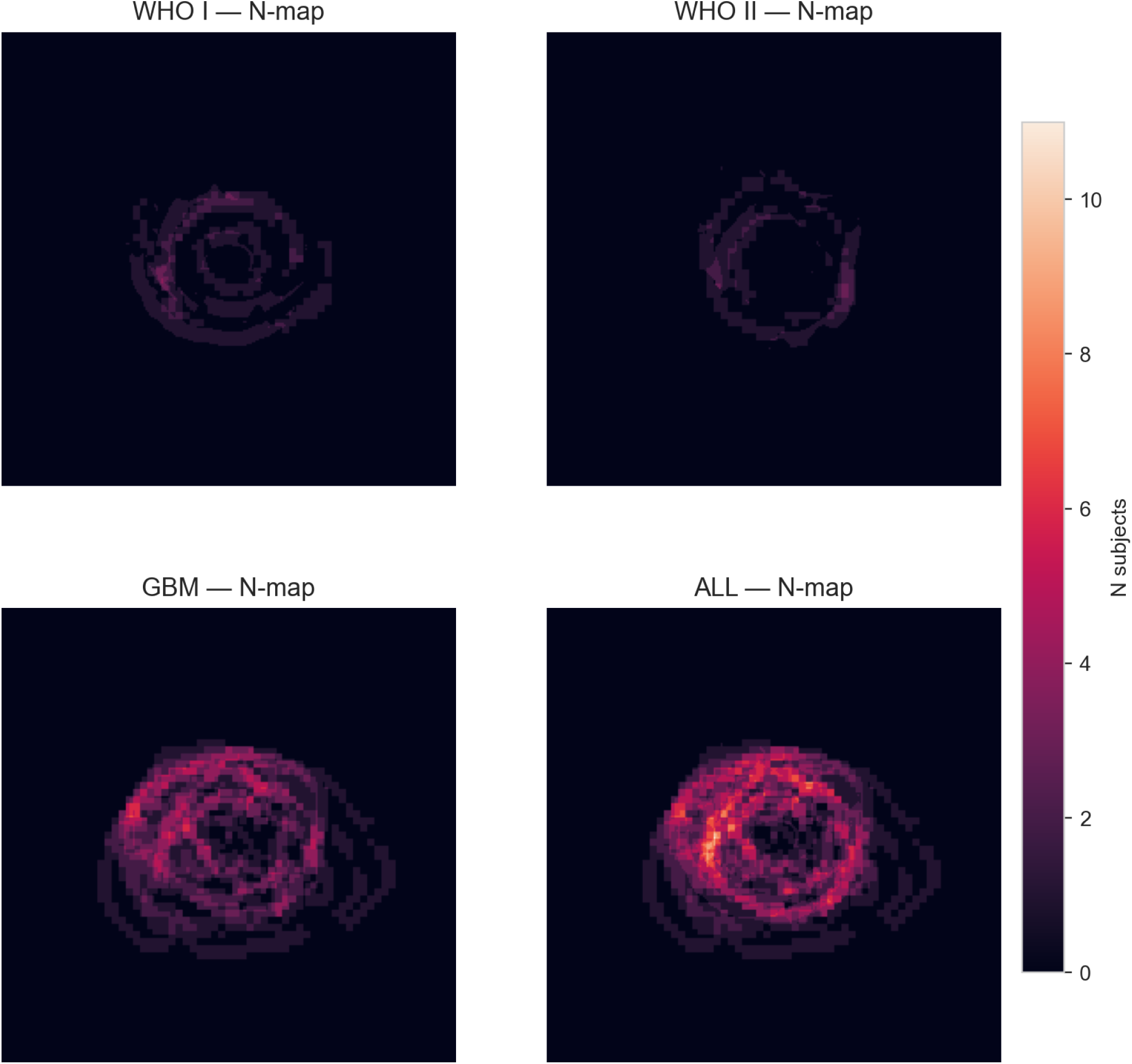
Group coverage (N-maps): Coverage maps showing the number of contributing subjects (N) per voxel after spatial normalization for WHO I, WHO II, GBM, and combined (ALL) cohorts. The maps visualize how many cases contained valid peritumoral tissue at each pixel, providing context for group-average reliability. Higher N values in the GBM and ALL maps reflect their larger sample sizes and broader peritumoral coverage.

## Notes

### Competing Interest Statement

The authors have declared no competing interest.

### Author Declarations

The study was conducted in accordance with the Declaration of Helsinki and approved by the Regional Committee on Health Research Ethics for the Region of Southern Denmark (IDs: S-20190105, S-20220055). All participants provided written informed consent prior to inclusion.

